# Perceptions of a Gender-Neutral Approach to Human Papillomavirus (HPV) Vaccination in Cameroon: A Qualitative Study

**DOI:** 10.1101/2025.04.29.25326670

**Authors:** Shariffatou Iliassu, Clarence Mbanga, Michael Budzi Ngenge, Shalom Ndoula, Andreas Ateke Njoh, Bridget C. Griffith, Sonali Patel, Navpreet Singh, Daniel Nebongo, Emilienne Carine Bieme-Ndi, Njike Sibenu Derrick, Shadrack Mngemane, Tosin Ajayi, Laure Anais Zultak, Yauba Saidu

## Abstract

Cameroon’s Expanded Program on Immunization (EPI) introduced the HPV vaccine into the routine immunization schedule in Cameroon in October 2020. However, HPV vaccine coverage has remained low, reaching only 20% by December 2022. In response, a gender-neutral vaccination (GNV) approach was adopted and implemented from January 2023 to increase acceptance, reduce stigma, and improve vaccine coverage. This study aimed to assess the perceptions of key immunization stakeholders in Cameroon on the GNV approach to HPV vaccination. A cross-sectional study was conducted from June to September 2024 using an in-depth and semi-structured interviews to explore the perspectives of key stakeholders (policymakers, community leaders, and parents of girls aged 9-13 years) across three regions (South-West, North, and Centre) and the central level. Data was analyzed using traditional content analysis, via manual and NVivo coding to identify key themes. The research team ensured reliability through consensus discussions, and key quotes illustrating central findings were identified. A total of 51 participants were interviewed. Participants observed that the inclusion of boys in the HPV vaccination program contributed to a positive shift in community perceptions, dispelled misconceptions, and enhanced overall vaccine acceptance. However, challenges related to limited resources, lingering misinformation, and hesitancy from parents, religious leaders, and healthcare workers hindered the effective implementation of the GNV approach. Participants emphasized the need for improved partnerships, sensitization strategies, integration with existing health programs, and sustained funding to enhance community understanding and increase vaccination uptake. Overall, stakeholders viewed the GNV approach as effective for improving HPV vaccine acceptability and promoting equity, but emphasized the need for continuous investments and stronger community engagement to address challenges like limited resources, misinformation, and hesitancy.

## 1. Introduction

Cervical cancer, primarily caused by persistent infection of high-risk strains of the human papillomavirus (HPV), particularly HPV types 16 and 18, is a public health challenge that has claimed the lives of about 350 000 women worldwide in 2022 [1,2]. In Cameroon, cervical cancer, with an estimated 2,525 new cases and 1,837 related deaths [3], ranks as the second most common cancer (after breast cancer) among women, highlighting the critical need for action to curb the burden associated with the disease.

In line with the significant contributions of cervical cancer to morbidity and mortality globally, the World Health Organization (WHO) developed a global strategy for the elimination of cervical cancer, centered around three key axes of intervention summarized in the 90–70–90 targets[4]. However, most low resource countries, Cameroon inclusive, have limited access to cervical cancer screening and treatment services, leaving HPV vaccination as the most cost-effective method to prevent cervical cancer and curb the burden linked to the condition in these settings. HPV vaccination, particularly when administered before sexual debut, has been shown to substantially reduce the risk of cervical cancer [5,6]. In line with this, Cameroon, via its Expanded Program on Immunization (EPI), introduced the HPV vaccine into its routine immunization calendar in October 2020, with a two-dose schedule for girls aged nine (09) years [7,8]. However, nearly two years post introduction, uptake has remained low, with coverage of 20% for the first dose recorded by the end of 2022, considerably lower than the 40% target established by the EPI for the introductory years of the vaccine [9].

A recent study in Cameroon identified several barriers to HPV vaccine uptake, including limited community awareness, insufficient use of media for education, and top-down health communication approaches[10]. Additional challenges include cultural misconceptions, inadequate stakeholder engagement, and hesitancy among healthcare workers due to concerns about vaccine safety, efficacy, and its perceived link to promoting promiscuity[8,11]. Furthermore, offering the HPV vaccination to girls only has perpetuated stigma and misinformation. [12,13]. To address these challenges, the country’s National Immunization Technical Advisory Group (NITAG) in June 2022 recommended transitioning to a single-dose HPV vaccination schedule, in alignment with guidance from the WHO Strategic Advisory Group of Experts on Immunization (SAGE)[14,15]. The NITAG also recommended a gender-neutral vaccination (GNV) approach to HPV vaccination, in a bid to foster inclusivity, increase acceptance, reduce stigma, and counter myths surrounding female-only vaccination.

Recommendations made by the NITAG were approved by the Ministry of Public Health and implemented as of January 2023, making Cameroon the first low– and middle-income country (LMIC) to implement the GNV approach for HPV vaccination. This was done alongside a revamped HPV vaccine delivery strategy involving community dialogues (CDs), followed by periodic intensification of routine immunization (PIRI) activities in 180 health areas across all ten regions of the country. This approach provides direct benefits such as reducing the risk of HPV-related diseases, and indirect benefits like subsequent protection for girls, [16,17] however, limited data exists on the effect GNV has on the perceptions and views surrounding the HPV vaccine in LMICs[18]. In addition, while GNV has been implemented and studied in high-income countries, there is limited data on its impact in LMICs[19].

This study aimed to fill the abovementioned gaps by assessing the perceptions, including awareness and the effect on HPV vaccine acceptability, of national and sub-national stakeholders on GNV implementation in Cameroon, to obtain insights required that could inform policy and practice around the successful implementation of the GNV approach in Cameroon and beyond.

## 2. Materials and Methods

### 2.1. Study design and duration

We conducted cross-sectional qualitative research over a four-month period between June to September 2024, to gain a more comprehensive and nuanced understanding of the perspectives of key national and sub-national stakeholders on the impact of the GNV approach to HPV vaccination awareness and acceptability in Cameroon.

### 2.2. Study Setting

This study was conducted in Cameroon, a country in the West-Central part of Africa that is administratively divided into ten regions (Adamawa, Centre, East, Far-North, Littoral, North, North-West, West, South and South-West) and has a population of approximately 30 million people in 2024[20]. The national official languages of the country are English and French. The delivery of healthcare in Cameroon is organized across three levels: central, intermediary (regional) and peripheral (district), each with defined responsibilities. The Ministry of Public Health, at the central level, is primarily responsible for policy formulation, strategic development, and system-wide coordination. The ten regional delegations serve as an intermediary layer, offering technical assistance to the 206 health districts where healthcare services are provided to the population.

Preventive services like immunization are integrated into all levels of the health system, with oversight on all vaccine introductions and vaccination activities provided by the central coordination of the Expanded Program on Immunization (EPI), and coordination at intermediate level ensured by regional arms of the EPI.

### 2.3. Study population and sampling

A convenience sampling approach was employed to select the study participants. Key informants at central and regional levels were purposively selected from the central and regional arms of the EPI respectively, based on their willingness to participate. At the district level, the health district lead (district medical officer) or their representative was selected for interviews. At the health area level, the EPI focal point or immunization staff from health facility harboring the health area coordinator (leading health facility) was selected. In addition, we identified and selected one traditional or religious leader per health area as a community leader to be interviewed. For parent representation, we purposively sampled four parents of girls aged 9-13 years in each of the 06x health areas, ensuring a mix of those who reported their daughters had and had not received the HPV vaccine.

To capture findings across different socio-economic strata and gain insights about varying contexts that would allow for a more nuanced understanding of the effectiveness of GNV across diverse settings, participants were selected from three regions (Southwest, North, and Centre) each offering unique contextual insights on HPV vaccination (South-west for conflict affected settings, North for rural populations with distinct cultural and economic contexts, and the Centre for urbanized areas). In each region, districts were stratified based on rural-urban status and the type of HPV intervention (GNV only or GNV+CD/PIRI), and two districts randomly selected per region as follows:

- Southwest: One urban GNV+CD/PIRI district and one GNV only rural district.
- North: One urban GNV only district and one rural GNV+CD/PIRI district.
- Centre: One urban GNV only district and one rural GNV+CD/PIRI district.

One health area was then randomly sampled from each selected health district. The sample selection process is summarized below (Fig 1).

**Fig 1:**
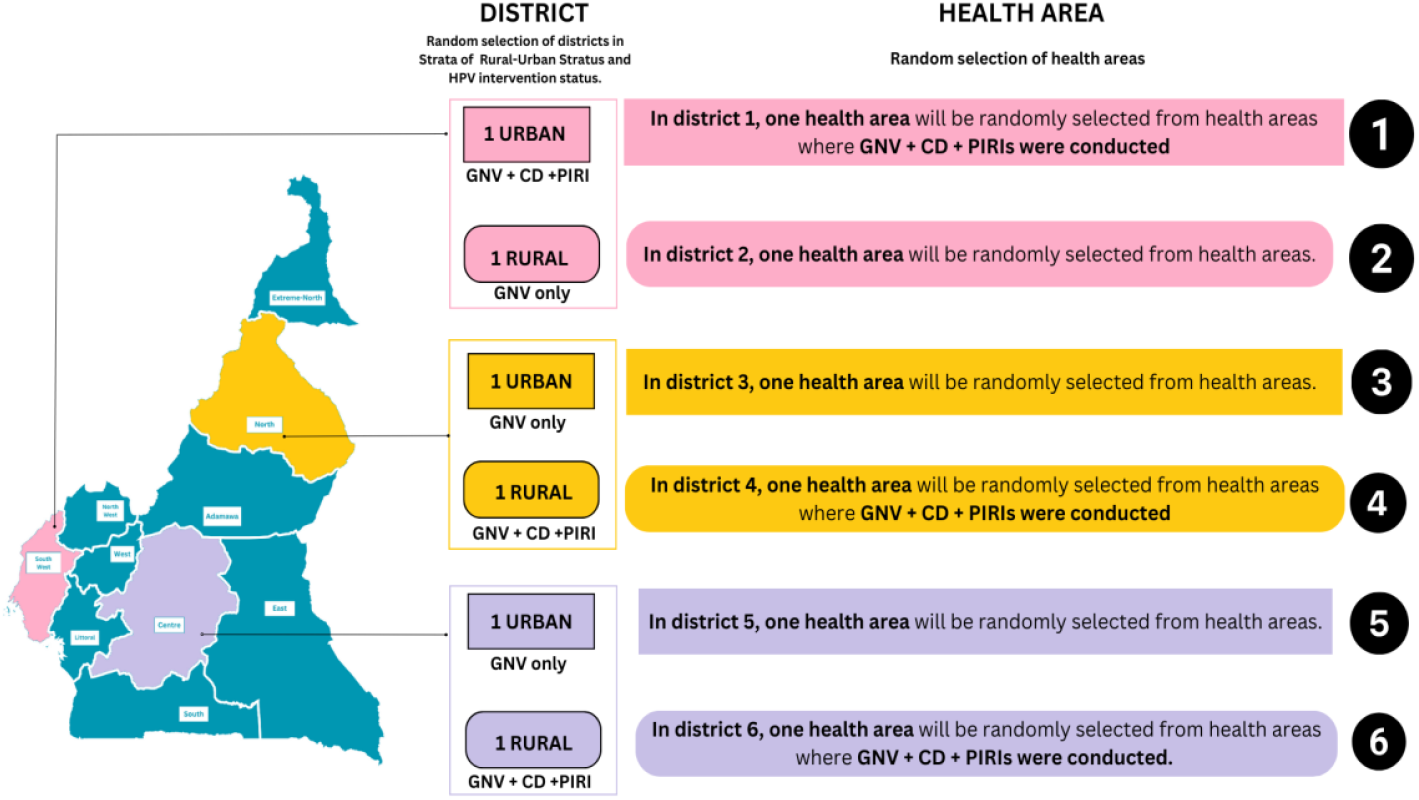
Sample selection process.

### 2.4. Data Collection

Data were collected via in-depth and semi-structured interviews with the different identified key stakeholders. In-depth interviews sought to assess stakeholders’ views and experiences of HPV vaccination, knowledge and perceptions of the GNV approach, barriers and facilitators of the GNV approach, and recommendations for the approach, while semi-structured interviews also obtained views and experiences on HPV vaccination, and the linking of CDs and PIRIs strategies to GNV. Prior to the interviews, selected key informants were contacted via email, phone or in person, the study’s purpose thoroughly explained to them, informed consent obtained and a suitable schedule for the interview agreed upon. All Key Informant Interviews (KIIs) were conducted in person by a trained data collector using an interview guide and lasted 30 to 45 minutes on average. Interviews were conducted in the preferred language of the participant and recorded for later transcription using an audio-recording device.

### 2.5. Data Management and analysis

Audio recordings from KIIs were transcribed verbatim, and all identifying information was removed with participants identified through secure coding systems. Access to the anonymized data was restricted to authorized study personnel with individual logins and role-based permissions. After data analysis, audio recordings were destroyed to protect the informants’ identities.

To analyze and triangulate the data, we employed traditional content analysis, a method chosen for its alignment with our study’s objectives and its effectiveness in analyzing interview data. Initial coding was based on both the interview guide and emerging themes from the data, resulting in three primary domains: effects of the GNV approach, challenges in implementation, and recommendations for adaptation. This coding was done using a combination of manual coding (to maintain a more flexible, grounded approach) and Nvivo software (to manage and organize the large volume of data). The Principal Investigator (PI), with input from the research team, extracted, compared, and categorized the codes, establishing connections between them and discussing these themes in relation to the study’s objectives during regular team meetings. Coding disagreements were resolved through structured team discussions, where all members worked to achieve consensus, with inter-coder reliability checks conducted to ensure consistency. In the final analysis phase, representative quotes were selected based on their illustrative power and ability to encapsulate key themes.

### 2.6. Ethical Consideration

This study was approved by Cameroon’s National Ethics Committee for Human Health Research (CNERSH) (application number 2024/06/168666/CE/CNERSH/SP). Informed consent was obtained from all participants involved in the study and their confidentiality and anonymity were maintained through unique identifiers, with no personal names or contact information used.

## 3. Results

### 3.1. Participant characteristics

A total of 51 Participants from three regions were interviewed. These participants were representatives of central level EPI (n=02), district representatives of the EPI (n=05), religious leaders (n=05), traditional leaders (n=04), and parents (n=24). More than half were from an urban health area (n=26). The distribution of study participants across the different study sites is outlined in Table 1 below.

**Table 1.**
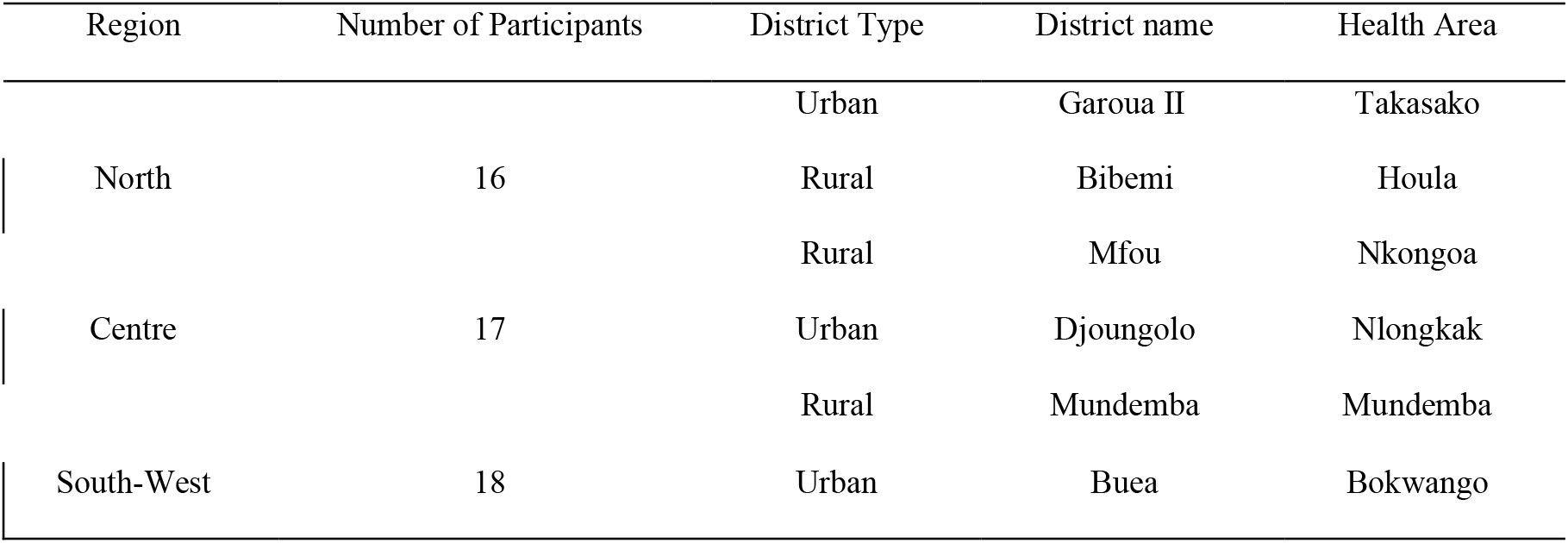
Distribution of study participants across the different study sites.

### 3.2 Themes

Three key themes related to the implementation of the GNV approach for HPV vaccination in Cameroon were identified across all stakeholder groups interviewed: (1) effects of GNV, (2) implementation challenges, and (3) adaptation strategies/recommendations.

#### 3.2.1 Effects of GNV

##### Increased Acceptability

Participants emphasized that incorporating both genders into the HPV vaccination program proved effective in addressing and countering prevalent myths and misconceptions, such as the incorrect belief that vaccinating girls might lead to in-fertility or other harm. By including boys in the vaccination effort, the program challenged these erroneous notions and promoted a more comprehensive understanding of the vaccine’s benefits. This inclusive strategy was perceived to significantly improved vaccine acceptance and reduced hesitancy among parents and caregivers.

*“Yes, certainly, since it can remove prejudices about the vaccine. In our community, we are used to vaccinating both boys and girls, so why are we now being asked to vaccinate only girls? That’s curious! If both sexes get vaccinated, it will make it easier for parents to accept the vaccine*…*”* **Parent, North region**

*“Yes here, there when people didn’t understand, but now it’s going to be fine. At the be-ginning, we said that the vaccine made you sterile, now people have understood how we have also integrated boys, it’s a matter that should help them. So, it’s the fact that it was a vaccine that sought to make girls sterile that is the problem*.” **Religious leader, North region**,

*“Yes. Now that they are giving it to both boys and girls, I believe the perception will really change, because there is this equality, and they know that it is not going to harm them. It’s not going to harm them, but it is for their own good*.*”* **Parent, South-West region**.

##### Increased Sense of Equity in Health Care Access

Participants highlighted that the implementation of GNV enhanced equity in healthcare by incorporating both boys and girls into the vaccination program. They felt that historically, boys were frequently excluded from such programs, despite the acknowledgment that HPV-related cancers also impact them.

*“Because there is this equality balance, this equality that it is not only for girl children, but for both sides. So, it will really boost the percentage. Since parents know that there is no discrimination between the two parties. And based on the explanation given to us by the doctor, at first it was only the girls that were given, and the boys were left alone. With the coming of both persons having it, it will encourage parents to go in, not to be neglected. Like, okay, me that all my children are boys, means that they have abandoned them*.*”* **Parent, South-West region**.

*“Yes, [the GNV approach] is a good idea. Even before the parents wondered why, just the girls. So, as now, there are also boys, it is good and will always help parents*.*”* **Parent, North region**.

*“Yeah, because if, like, it is only for the girl child, I think the work is incomplete. Now that it is administered to both sexes this is better, I think it is ok and should continue*. **Parent, North region**.

##### Increased Social Cohesion

Redirecting the focus from vaccinating a particular gender to including all children promoted social cohesion and a collective sense of responsibility within the community. Many stakeholders believed this strategy effectively increased community engagement and participation in the vaccination initiative.

*“But yes, medicine has evolved a lot today. So, knowing that we vaccinate men makes me very happy and it warns us of a lot of diseases in our children. So, I’m for those who vaccinate, whether it’s women or men, I encourage them to do it*…*”* **Parent, Centre region**.

*“No, no, it’s a very good initiative and it comes at the right time, knowing that health is a priority and a concern for parents. It would be very important if more of such opportunities were created. Because really neither boys nor girls are safe from these different pathologies….“* **Parent, Centre region**.

#### 3.2.2 Implementation challenges of the GNV approach

##### Resource Gaps

Despite its successes, the implementation encountered bottlenecks related to human and material resources, including staff shortages and increased costs associated with vaccinating boys. Representatives from the EPI at both the central and district levels emphasized that the additional financial burden affected logistics (transportation, vaccination equipment, vaccines, tokens for community leaders and personnel (health facility staff, staff training, community relay agents)), often necessitating out-of-pocket expenditures to implement the GNV strategy. However, this approach is not sustainable, highlighting the urgent need for increased funding and staffing

*“Logistics, transportation, per diem, insurance, personnel training and equipment, vaccination products, tokens for community leads for their involvement, are resources which need to be deployed for an effective implementation of vaccination projects. However, they are often not reimbursed or budgeted for. This constitutes a financial burden on health facilities which in most cases can’t cover all the cost incurred to reach all the target population”* **EPI representative, central leve**l

*“The other challenge is that we also have a problem of staff and human resources. Because in some health areas…the staffing there is extremely low. And with the situation that we are having now, the crisis, and also the PBF has completely disappeared, so the autonomy of health facilities in terms of financial resources is completely lost. Those are the challenges*.*”* **EPI representative, South-West region**.

*“Yes, because normally in order to succeed, that is to say good awareness, as you said, you have to go and meet the parents to ask, that is to say to make them aware, to tell them, to convince them of the importance of the vaccine against the virus. It requires financial means, logistical means and everything, but no means have been made available to us to be able to overcome these challenges*.*”* **EPI representative, North region**”

##### Limited Community Awareness on HPV vaccination and the GNV approach

Many parents and community members mentioned they had insufficient awareness or understanding of the HPV vaccination, and the GNV approach, often resulting from missed opportunities for sensitization and ineffective communication strategies.

*“Personally, I didn’t find out that there were [HPV vaccine] awareness-raising activities in Takasko, especially as I’m close to the chiefdom. The people in charge of information didn’t give out the information I needed. I didn’t hear anything, otherwise my children would have been vaccinated*.*”* **Parent, North region**.

*“I didn’t have my children vaccinated [for HPV], since I’m not even aware of the vaccination”* **Traditional leader, North region**

*“Well for the boys I wasn’t really aware of it, but for the girls I know very well*.” **Religious leader, Centre region**.

*“I think I am hearing this [boys being vaccinated] for the first time*.*”* **Parent, South-west region**.

##### Misinformation

The prevalence of misinformation on social media, particularly from anti-vaccination activism, necessitates pro-active measures to combat false information and promote accurate details about the HPV vaccine’s benefits. Parents mentioned that while there is a general willingness to vaccinate within the community, specific fears related to the HPV vaccine amplified by misinformation, have discouraged parents from seeking vaccination. Exposure to social media videos claiming the vaccine aims to reduce the African population and could cause infertility has instilled fear and hesitation among parents. Overall, despite GNV normalising the HPV vaccine, significant challenges remain in Cameroon due to ongoing misinformation, especially on social media

*“I wanted to give my children the vaccine but later they sent us some videos on social media that the white people want to reduce the African population by making sure they don’t give birth again. That if you give that vaccine to your daughter, she will not give birth. So, that’s the reason I did not give my daughter the vaccine because I equally received the videos and was afraid that I can give my daughter the vaccine and she never put to birth*.*”* **Parent, South-West region**.

*“When I arrived, I discovered that there was reluctance among religious leaders who did not agree with this vaccine, supposedly that it made girls sterile*.*”* **EPI representative, North region**.

*“Even if others let their children get vaccinated, there is even more resistance to this vaccine, because of the misinformation circulating about the vaccine*.*”* **EPI representative, North region**.

##### Hesitancy

###### Parents

Parents consistently reported hesitancy toward vaccinating their children, both at the individual and community levels. This hesitancy was driven by fears of potential side effects, widespread myths and misinformation such as concerns about infertility and social dynamics, including the need for paternal consent. These factors significantly influenced decision-making around HPV vaccination, contributing to delayed uptake or refusal.

*It’s because, you know when it concerns vaccination many of us were afraid you understand. So, when this vaccine came up I had not given it to my child before so normally I must be afraid because I don’t know the effect of that vaccine. My child may be injected now and maybe it will affect one of his hands or leg, so I had doubts*. **Parent, South-west region**.

*“She hasn’t received the vaccine yet*..*I am still hesitant on this subject*… *I didn’t want that no no I hesitated because of the problems of vaccines and everything we hear about vaccines…And then her father has to give his approval. Her father must say…Because the community is a little stuck if we can say so. Everyone is hesitant. Everyone is afraid. People are afraid of vaccines*.*”* **Parent, Centre region**.

*“Many have heard here and there that vaccines cause diseases, that they will make girls sterile, prevent boys from having children later on, that’s why they have stuck to their positions and not vaccinating*.” **Parent, North region**

###### Health Workers

Representatives from the EPI noted that initial hesitancy toward HPV vaccines for girls was mostly observed among health workers who lacked adequate information about HPV vaccination from central to peripheral levels. This information gap contributed to skepticism and distrust surrounding the vaccine. Health care workers who expressed distrust and hesitancy reflected this in their work and spread it to the communities. To address this issue, the EPI reinforced communications on HPV vaccination and training initiatives for health staff.

*“Yeah, health workers were fueling the resistance, that’s the truth*…*You know health workers who are not in immunization have as little information as the rest of the public*… *so, they were suspicious at their own end, a lot of people even within the health system in Cameroon did not trust the process*… *you can even see in terms of the willingness of vaccination health facilities to request HPV vaccines*.*”* **EPI representative, Central level**.

*“We realized that there was a paucity of information on the side of our colleagues who work in the health facilities. So, it is our responsibility to inform them properly, to sensitize them and since they have started having the right information, you can see more willingness on the part of health workers to get involved”* **EPI representative, Central level**.

*“Yes, the first barrier to the implementation of this vaccination is first and foremost the health personnel. You meet your colleagues who have the target children, who hesitate, who think twenty times, who are even afraid, who sometimes even say you wonder if they really deserve to be healthy. Well now the other resistance is social media, the church as well*.*”* **EPI representative, Centre region**.

###### Religious leaders

There were noticeable religious barriers to HPV vaccination efforts, highlighting how certain faith leaders can hinder community acceptance and participation. Participants explained that many individuals are influenced by their religious leaders, who may discourage vaccination based on their beliefs. This resistance is compounded by the lack of cooperation from religious health facilities, which can act as important communication channels for health initiatives. For instance, in some cases, priests refused to allow health representatives to address congregations or distribute informational materials, necessitating intervention from local authorities to facilitate discussions.

*“Sometimes it’s related to their religions. There are sometimes others who feel that their religions do not allow them to be vaccinated*.*”* **Traditional leader, Centre region**.

*“Religious leaders for HPV… I’m a Catholic, but I would say… our priest was not too welcoming with the HPV. When we sent a…, well, he didn’t even attend the advocacy meeting. I personally went there to like try to educate and then pleaded with him to give me, like five minutes during the church service to just say something. He was like…, no, if the parents want to give their children, they should give, but it will not be done… I will not give you that time to say, sorry to say that. I will not. So, slowly we are still working on convincing…”* **EPI representative, South-west region**.

*“When I arrived, I discovered that there was reluctance among religious leaders who did not agree with this vaccine, supposedly that it made girls sterile*.*”* **EPI representative, North region**.

#### 3.2.3 Key adaptation strategies/Recommendations

##### Partnerships and collaboration

Participants universally conveyed that a collective approach wherein each stakeholder feels valued and informed, significantly boosted community engagement and enhanced the GNV approach and HPV vaccination coverage. They depicted collaboration as a multi-faceted effort that not only involved health workers but also extended to educational authorities and religious leaders, illustrating that shared communication and joint efforts were essential for effective outreach activities especially. Participants noted that cross-sector engagement fostered a supportive environment for sensitization and consent, ultimately building trust and encouraging HPV vaccine acceptance among families. As such, they advocated higher-level directives to strengthen these partnerships.

*“… I say to myself at the ministerial level, if between the Minister of Health and the Minister of Basic Education perhaps, if the instruction is given there where each minister tells his collaborators that no, here is what we have to do, it can also be easy, because when we leave at our level here, And I’m going to see the inspector there, it can’t work. But if we tell them, an instruction comes from the hierarchy where we tell them that no, here is the partnership, the collaboration, there it can work. that the instruction of collaboration be given a little higher”* **EPI representative, South-west region**.

*“Always use leaders and community agents, these groups will be very good for the propaganda and effectiveness of this GNV approach*.*”* **Religious leader, North region**.

*“We have to continue to work well*… *We [leaders] work well with the hospital. If there is something to do, let him summon us, let him tell us*.*”* **Traditional leader, North region**.

*“So, to facilitate access to the vaccine, in the community, you have to take people, how can I put it? Uh, leaders of the neighborhood who have a good image and who have the ease of communication with the population, so, they will make it easier for you to vaccinate*.*”* **Parent, Centre region**.

##### Sensitization of health providers and communities

Participants reported community engagement was essential, as empowering local leaders both traditional and religious enabled them to drive awareness campaigns and disseminate accurate information. They ultimately recommended that by intensifying educational activities and fostering a deeper understanding of HPV and its preventive measures, communities are more likely to embrace vaccination.

*“Yes, eh in relation to the non-gender strategy eh… capacity building was ensured uh in the coordination meetings*…, *we strengthened the capacities of community relays, we strengthened the capacities of traditional and religious leaders and uh here it is… We often use them to drive mini campaigns in communities. So, when we have a community to raise awareness, we bring them down to implement the activity*.*”* **EPI representative, Centre region**.

*“Simply raising parents’ awareness of the importance of vaccination and providing them with clear, precise information on the benefits of vaccination could dispel the fears and false beliefs that surround vaccines*.*”* **Parent, North region**.

##### Integration with existing health programs

Participants from the EPI and community level reported that integrating the HPV vaccination into established health programs, such as polio vaccination campaigns, significantly enhanced both acceptance and uptake of the vaccine. This approach leveraged the existing infrastructure and familiarity of ongoing health interventions, creating a seamless experience for the target population and facilitating a smoother introduction and administration process.

*“While we are conducting our home visits or we are doing a vaccination campaign, we still introduce HPV during the briefings, we already have when we have a vaccination campaign you will always see the people lost to follow-up with such a vaccine, and it’s not formal, it’s not official but we still ask the community health workers who go to the field if you have met anyone to whom you can talk about HPV, you talk to him*.*”* **EPI representative, Centre region**.

*“The advantages are that they already know about other vaccines like malaria, measles and others. This favors acceptance of other vaccines, especially HPV, and the strategy used is continuous awareness-raising*.*”* **Traditional leader, North region**.

## 4. Discussion

To improve HPV vaccination acceptance and coverage, Cameroon adopted a GNV approach to HPV vaccination in 2023. This study explored the perceptions of key stakeholders across different tiers of the health system on this approach, in a bid to generate insights that could inform policy and practice GNV implementation in low resource settings. Overall, we found that the inclusion of boys in HPV vaccination challenged myths about HPV vaccination, resulting in a positive shift in community perceptions about the vaccine and improved acceptance. However, challenges persist particularly in resource allocation, community awareness and lingering misinformation.

The introduction of a GNV strategy was recognized by stakeholders as an effective approach to improve acceptability and HPV vaccine coverage among both girls and boys. This underscores the potential of inclusive health interventions that address gender equity in healthcare access, as outlined in the Immunization Agenda 2030, which acknowledges that HPV-related cancers affect both genders[21,22] [20,21]. It also supports the conclusions from a recent systematic review which advocates for a GNV approach, noting that a girls-only strategy could not only be discriminatory but could also foster confusion and reinforce gender inequalities[23,24]. The perception of increased acceptability in our study can be attributed to a multifaceted approach that included community engagement and strategic stakeholder collaboration, reinforcing the positive impact of comprehensive health campaigns. Moreover, including boys in the vaccination program served as a pivotal step in combating myths that fueled vaccine hesitancy, such as the erroneous belief that vaccination could result in infertility for girls. By broadening the focus of HPV vaccination beyond girls, the program reduced gender-specific skepticism, promoting a more informed and balanced understanding of HPV prevention. Several studies have highlighted diverse benefits of extending the HPV vaccine to boys, including decreased prevalence of anogenital cancers among males, and herd-immunity[25,26]. However, our study brings forward a new benefit, which is the reduction of myths around the vaccine and increased acceptance in LMICs. This finding has implications for future vaccination strategies, suggesting that gender neutrality can be an effective means of mitigating unfounded health concerns and fostering community trust.

Despite the impact of the GNV approach, the HPV vaccination program in Cameroon faced notable obstacles that need to be addressed to sustain and improve its outcomes. A critical issue was the scarcity of resources, including staff shortages and rising operational costs. These financial and logistical challenges placed significant strain on health facilities, often forcing them to rely on out-of-pocket expenses for continued service delivery. Although our study did not assess cost-effectiveness, it highlights the real-world operational constraints that can influence the feasibility and impact of the GNV approach in resource-limited settings. The lack of reimbursement for essential costs, such as personnel training, transportation, and community incentives, underscores the pressing need for secure funding models to support vaccination programs.

Ongoing vaccine hesitancy was also noted among healthcare workers, especially those who lacked information on immunization. Their limited knowledge about the HPV vaccine contributed to wariness, which was evident in their interactions with communities. This observation is echoed in findings from two recent systematic reviews which showed that healthcare workers were less inclined to recommend or promote vaccination when they had concerns about the vaccine’s safety and efficacy, coupled with insufficient knowledge and awareness. To address this issue, targeted training and capacity-building initiatives are essential to bolster healthcare workers’ confidence and enable them to effectively advocate for the vaccine [27,28].

The study highlighted several ways to address these barriers and enhance the HPV vaccination program and the GNV approach. Partnerships and cross-sector collaboration were recognized as essential for building trust and boosting the effectiveness of vaccination campaigns, aligning with findings from other research [18,29]. Involving community leaders, including traditional and religious figures, can improve message acceptance and encourage participation, even in the face of initial resistance.

Educational and sensitization efforts were deemed necessary for improving understanding among both health providers and the public. Tailored training programs for health workers can mitigate skepticism and ensure they can confidently inform and promote vaccination. Also, empowering community leaders to drive localized awareness campaigns helps dispel myths and provide accurate information, thus reducing vaccine hesitancy. Similar findings are captured in various studies which reveal that awareness raising at various levels as well as higher knowledge of health care workers on HPV vaccination increased vaccine confidence and acceptance rates [18,30].

Integration of HPV vaccination into existing health programs, was outlined as an important mechanism to improve vaccine acceptance by leveraging established trust and infrastructure. This approach decreases the novelty and logistical burden of new vaccination efforts, facilitating smoother implementation and higher community participation. However, there was still a lack of continuous financial support that is important for overcoming resource constraints, a similar pattern observed in studies carried out in various lower– and middle-income countries [31–34]. The study advocates for dedicated funding that supports the full range of necessary resources, including transportation, training, community mobilization, and incentives for local leaders and participants in outreach activities.

## 5. Conclusions

Stakeholders perceived the GNV approach as effective in improving HPV vaccine acceptability among both girls and boys, shifting community perceptions, reducing stigma and addressing gender disparities in healthcare access in Cameroon. However, barriers related to resources, lingering misinformation, hesitancy, and logistics must be strategically addressed to sustain and scale this initiative. Collaborative partnerships, targeted education, and integrated health approaches are essential to overcoming these barriers. Future steps should focus on securing long-term funding and resource allocation to make the HPV vaccination program and the GNV approach a sustainable and effective component of public health policy.

## Data Availability

Data used for this research are available on CHAI – Cameroon Box Folder at https://clintonhealth.app.box.com/folder/280231441720

## Acknowledgments

The authors would like to express their sincere appreciation for the invaluable support provided by the members of the Central and Regional EPI teams, as well as the leadership of the respective target health districts. Their facilitation, coordination, and commitment were essential in ensuring the smooth implementation of this study across the selected regions and districts. Furthermore, we extend our deepest gratitude to the healthcare facility staff for their collaboration and dedication, as well as to the community members (parents, religious leaders, and traditional leaders) whose willingness to participate and share their insights was instrumental in shaping the findings of this research. Their collective contributions were fundamental to the successful completion of this study and to the generation of meaningful evidence that can inform GNV implementation for HPV vaccination.

## References

1. Okunade KS. Human Papillomavirus and Cervical Cancer. J Obstet Gynaecol J Inst Obstet Gynaecol. 2020 Jul;40(5):602–8.

2. Cervical cancer [Internet]. [cited 2025 Mar 11]. Available from: https://www.who.int/news-room/fact-sheets/de-tail/cervical-cancer

3. Cancer Today [Internet]. [cited 2024 Dec 18]. Available from: https://gco.iarc.who.int/today/

4. Global strategy to accelerate the elimination of cervical cancer as a public health problem [Internet]. [cited 2024 Dec 18]. Available from: https://www.who.int/publications/i/item/9789240014107

5. Kamolratanakul S, Pitisuttithum P. Human Papillomavirus Vaccine Efficacy and Effectiveness against Cancer. Vaccines. 2021 Dec;9(12):1413.

6. Rylander E, Ruusuvaara L, Almströmer MW, Evander M, Wadell G. The absence of vaginal human papillomavirus 16 DNA in women who have not experienced sexual intercourse. Obstet Gynecol. 1994 May;83(5 Pt 1):735– 7.

7. McLemore MR. Gardasil: Introducing the new human papillomavirus vaccine. Clin J Oncol Nurs. 2006 Oct;10(5):559–60.

8. Ogembo JG, Manga S, Nulah K, Foglabenchi LH, Perlman S, Wamai RG, et al. Achieving high uptake of human papillomavirus vaccine in Cameroon: Lessons learned in overcoming challenges. Vaccine. 2014 Jul 31;32(35):4399–403.

9. Ministry of Public Health. DHIS2 immunisation portal [Internet]. 2022. Available from: https://dhis-minsante-cm.org/dhis-web-data-visualizer/index.html

10. Elit L, Ngalla C, Afugchwi GM, Tum E, Domgue JF, Nouvet E. Assessing knowledge, attitudes and belief toward HPV vaccination of parents with children aged 9–14 years in rural communities of Northwest Cameroon: a qualitative study. BMJ Open. 2022 Nov 15;12(11):e068212.

11. Haddison E, Tambasho A, Kouamen G, Ngwafor R. Vaccinators’ Perception of HPV Vaccination in the Saa Health District of Cameroon. Front Public Health [Internet]. 2022 Jan 10 [cited 2024 Dec 16];9. Available from: https://www.frontiersin.org/journals/public-health/articles/10.3389/fpubh.2021.748910/full

12. Khosa LA, Meyer JC, Motshwane FMM, Dochez C, Burnett RJ. Vaccine Hesitancy Drives Low Human Papillomavirus Vaccination Coverage in Girls Attending Public Schools in South Africa. Front Public Health. 2022 May 24;10:860809.

13. Pappano H. Women and Vaccines: Bridging the Gap for Gender Equity [Internet]. Shot@Life.2024 [cited 2025 Mar 14]. Available from: https://shotatlife.org/2024/10/10/women-and-vaccines/

14. Strategic Advisory Group of Experts on Immunization (SAGE) – April 2022 [Internet]. [cited 2024 Dec 18]. Available from: https://www.who.int/news-room/events/detail/2022/04/04/default-calendar/sage_meeting_april_2022

15. Note de Recommandation n° 009/2022 du Groupe Technique Consultatif National pour la Vaccination (GTCNV) A/s Priorité de recherche opérationnelle sur la vaccination anti-HPV. 2022.

16. Bogaards JA, Wallinga J, Brakenhoff RH, Meijer CJLM, Berkhof J. Direct benefit of vaccinating boys along with girls against oncogenic human papillomavirus: bayesian evidence synthesis. BMJ. 2015 May 12;350:h2016.

17. Marty R, Roze S, Bresse X, Largeron N, Smith-Palmer J. Estimating the clinical benefits of vaccinating boys and girls against HPV-related diseases in Europe. BMC Cancer. 2013 Jan 8;13:10.

18. Yangchen S, Felsher M, Acosta D, Sukarom I, Wu L, Phuntsho S, et al. Lessons Learned From Bhutan on extending Girls-Only HPV Vaccination Program to Boys: A Qualitative Study. Asia Pac J Public Health. 2024 Sep;b36(6– 7):580–8.

19. Griffith BC, Iliassu S, Mbanga C, Budzi MN, Patel S, Singh N, et al. Assessing the impact of a gender-neutral approach to HPV vaccination on vaccination coverage for nine-year-old girls in Cameroon.

20. Cameroon population 2024 – StatisticsTimes.com [Internet]. [cited 2025 Apr 4]. Available from: https://statisticstimes.com/demographics/country/cameroon-population.php

21. Immunization Agenda 2030 [Internet]. [cited 2024 Dec 12]. Available from: https://www.who.int/teams/immun-ization-vaccines-and-biologicals/strategies/ia2030

22. Giuliano AR, Nyitray AG, Kreimer AR, Pierce Campbell CM, Goodman MT, Sudenga SL, et al. EUROGIN 2014 roadmap: Differences in human papillomavirus infection natural history, transmission and human papillomavirus-related cancer incidence by gender and anatomic site of infection. Int J Cancer. 2015;136(12):2752–60.

23. Dykens JA, Peterson CE, Holt HK, Harper DM. Gender neutral HPV vaccination programs: Reconsidering policies to expand cancer prevention globally. Front Public Health. 2023 Feb 21;11:1067299.

24. Deignan C, Swartz A, Cooper S, Colvin CJ. Stakeholders’ Understandings of Human Papillomavirus (HPV) Vaccination in Sub-Saharan Africa: A Rapid Qualitative Systematic Review. Vaccines. 2021 May 12;9(5):496.

25. Majed L, Bresse X, El Mouaddin N, Schmidt A, Daniels VJ, Pavelyev A, et al. Public health impact and cost-effectiveness of a nine-valent gender-neutral HPV vaccination program in France. Vaccine. 2021 Jan 8;39(2):438– 46.

26. HTA of extending the HPV vaccination to boys | HIQA [Internet]. [cited 2024 Dec 12]. Available from: https://www.hiqa.ie/reports-and-publications/health-technology-assessment/hta-extending-hpv-vaccination-boys

27. Osaghae I, Darkoh C, Chido-Amajuoyi OG, Chan W, Padgett Wermuth P, Pande M, et al. Healthcare Provider’s Perceived Self-Efficacy in HPV Vaccination Hesitancy Counseling and HPV Vaccination Acceptance. Vaccines. 2023 Jan 30;11(2):300.

28. Lin C, Mullen J, Smith D, Kotarba M, Kaplan SJ, Tu P. Healthcare Providers’ Vaccine Perceptions, Hesitancy, and Recommendation to Patients: A Systematic Review. Vaccines. 2021 Jul;9(7):713.

29. Gallagher KE, Howard N, Kabakama S, Mounier-Jack S, Burchett HED, LaMontagne DS, et al. Human papillomavirus (HPV) vaccine coverage achievements in low and middle-income countries 2007–2016. Papillomavirus Res. 2017 Dec 1;4:72–8.

30. Bakare D, Gobbo E, Akinsola KO, Bakare AA, Salako J, Hanson C, et al. Healthcare worker practices for HPV vaccine recommendation: A systematic review and meta-analysis. Hum Vaccines Immunother. 2024 Dec 31;20(1):2402122.

31. Ladner J, Besson MH, Hampshire R, Tapert L, Chirenje M, Saba J. Assessment of eight HPV vaccination programs implemented in lowest income countries. BMC Public Health. 2012 May 23;12:370.

32. Wigle J, Coast E, Watson-Jones D. Human papillomavirus (HPV) vaccine implementation in low and middle-income countries (LMICs): Health system experiences and prospects. Vaccine. 2013 Aug 20;31(37):3811–7.

33. Hutubessy R, Levin A, Wang S, Morgan W, Ally M, John T, et al. A case study using the United Republic of Tanzania: costing nationwide HPV vaccine delivery using the WHO Cervical Cancer Prevention and Control Costing Tool. BMC Med. 2012 Nov 13;10:136.

34. Ladner J, Besson MH, Audureau E, Rodrigues M, Saba J. Experiences and lessons learned from 29 HPV vaccination programs implemented in 19 low and middle-income countries, 2009–2014. BMC Health Serv Res. 2016 Oct 13;16:575.

